# Mathematical Mo Del with So Cial Distancing Parameter for Early Estimation of Covid-19 Spread

**DOI:** 10.1101/2020.04.30.20086611

**Authors:** Saroj Kumar Chandra, Avaneesh Singh, Manish Kumar Bajpai

## Abstract

COVID-19 is well known to everyone in the world. It has spread around the world. No vaccine or antiviral treatment is available till now. COVID-19 patients are increasing day by day. All countries have adopted social distancing as a preventive measure to reduce spread. It becomes necessary to estimate the number of peoples going to be affected with COVID-19 in advance so that necessary arrangements can be done. Mathematical models are used to provide early disease estimation based on limited parameters. In the present manuscript, a novel mathematical model with a social distancing parameter has been proposed to provide early COVID-19 spread estimation. The model has been validated with real data set. It has been observed that the proposed model is more accurate in spread estimation.

## 1. Introduction

COVID-19 has emerged as a life-threatening outbreak disease. World Health Organization has declared as a pandemic in January 2020^1^. The first case has been reported in Wuhan city of Hubei Province in south China on 31, December 2019 as unidentified pneumonia^2, 3^. COVID-19 disease has been identified as a member of the Severe Acute Respiratory Syndrome (SARS) that outbroke also in South China in 2002-2003^4^. This disease is developed in the human body in the presence of a coronavirus. Tyrell and Bynoe have described the coronavirus in 1966^5^. The virus is divided into four categories namely alpha, beta, gamma, and delta. Bats are a major source of alpha and beta virus category of the coronavirus. While gamma and delta originate from pigs and birds. Among these viruses, beta can infect human beings. The most common symptoms of COVID-19 are fever, tiredness, and dry cough. Some patients may have aches and pains, nasal congestion, runny nose, sore throat or diarrhea. Around 1 out of every 6 people who get COVID-19 becomes seriously ill and develops difficulty breathing^6^. COVID-19 is a transmissible disease. Hence, peoples get infected with coronavirus from others (COVID patients) by coughs or sneezing^7^. No vaccine or antiviral treatment is available until now. The mortality rate is increasing day by day. The rapid spread of the COVID-19 may be due to multiple causes. One cause is the lacking of information transparency at the early stage of the epidemic outbreak. Releasing the epidemic information in a timely and accurate way is extremely important for the anti-epidemic response of the public. The authentic and transparent information could have prohibited the spread of the COVID-19 at the early stage. The other cause is the lacking of the scientific diagnostic criterion for the COVID-19. As preventive measures, all the major countries have canceled events, classes in schools and colleges, businesses have pushed work from home policies. All of these measures are adopted to slow the spread of the disease. These measures are broadly referred to as social distancing. In the present manuscript, a mathematical model is being developed for estimating COVID-19 disease. In addition, the proposed model incorporates the social distancing parameter for better spread estimation.

Mathematical modelling has gained more attention and awareness in epidemiology and the medical sciences^8-11^. It has been used for cancer detection, segmentation and classification^12, 13^. These models are useful in cases where disease dynamics are not unclear. It estimates the number of cases in worst and best-case scenarios. Susceptible-Infected-Removed (SIR) and Susceptible-Exposed-Infected-Removed (SEIR) models are the most studied and used models to estimate disease spread. These models are helpful in estimating the effect of preventive measures on disease spread. The SIR model originated from the study of the plague almost one hundred years ago. This model estimates spread by using contact rate and infection period only. It has been observed that this model is not suitable for estimating the spread in which the incubation period is involved such as COVID-19. Hence, the SEIR model is a suitable model for estimating COVID-19 spread since it incorporates the incubation period. However, the SEIR model failed to estimate the spread where preventive measures are adopted such as social distancing. In the present manuscript, a modified SEIR model is presented estimating COVID-19 spread. The proposed model has incorporated the social distancing parameter more accurate estimation of COVID-19 disease.

The present work is organized as follows. The mathematical model for COVID-19 spread estimation is presented in section 2. Section 3 discusses the results obtained by the proposed mathematical model.

## 2. Proposed Methodology with Social Distancing

In this section, a modified SEIR mathematical model is being presented for estimating COVID-19 spread. Proposed model uses basic SEIR model to design novel mathematical model. The SEIR model is a compartmental model for estimating disease spread in the population. It’s an acronym for Susceptible, Exposed, Infected, Recovered. When a disease is introduced to a population, the people move from one compartment to another compartment. When a people has reached to R state, then the people either survived the disease or out of the population. The classical SEIR is defined be equations

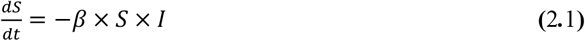

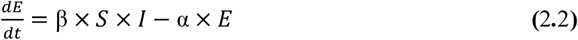

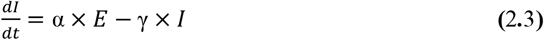

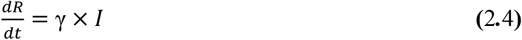

Here, Eq. (2.1), (2.2), (2.3) and (2.4) represent four Ordinary Differential Equations (ODE’s). The three parameters are α, β and γ. These are defined as

α is the inverse of the incubation period

β is the average contact rate in the population

γ is the inverse of the mean infectious period

Eq. (2.1) is the change in people susceptible to the disease and is moderated by the number of infected people and their contact with the infected. Eq. (2.2) gives the people who have been exposed to the disease. It grows based on the contact rate and decreases based on the incubation period whereby people then become infected. Eq. (2.3) gives us the change in infected people based on the exposed population and the incubation period. It decreases based on the infectious period, so the higher γ is, the more quickly people die/recover and move on to the final stage in Eq. (4). The proposed mathematical model is shown in the Fig. 1

**Fig. 1.**
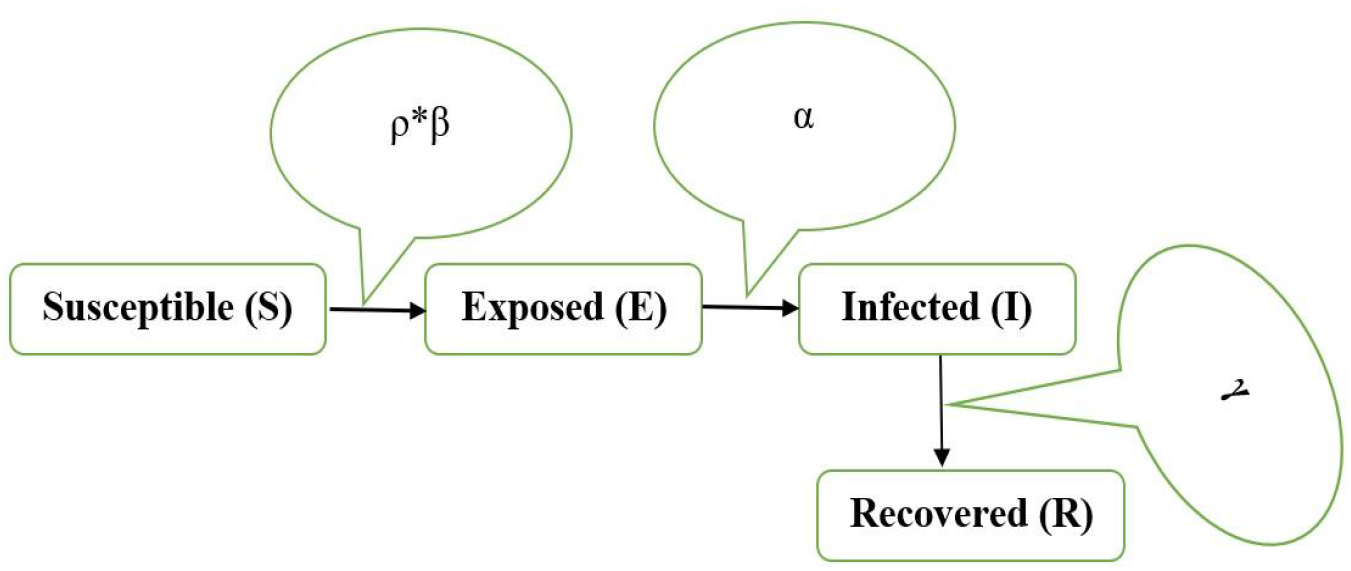
Proposed Mathematical Model with Social Distancing

In the proposed model a novel factor ρ has been introduced as social distancing parameter. Social distancing is maintained in transmissible diseases like COVID-19. It includes avoiding large gatherings, physical contact, and other efforts to mitigate the spread of infectious disease. This parameter controls contact rate, β. Social distancing parameter ρ lies in the range 0 and 1, where 0 indicates everyone is locked down and quarantined while 1 is equivalent to our base case above. The Eq. (2.1) and (2.2) are being multiplied with parameter to have social distancing effect. The modified equations are defined as

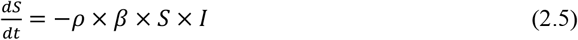

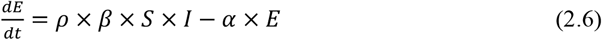

## 3. Results and Discussion

All the experimental studies have been performed on PYTHON. The machine used for performing simulation work has CPU clock speed 1.60 GHz, 8GB RAM, 256 KB L1 cache, 1.0 MB L2 cache, and 6.0 MB L3 cache hardware configuration. The proposed mathematical model has been validated on cases of COVID-19 in India^14^. The day-wise COVID-19 cases are shown in the Fig.2 State-wise active COVID-19 cases in India is shown in Fig. 2^15^. It can be analyzed from Fig.2 that COVID-19 cases are increasing day by day.

**Fig. 2.**
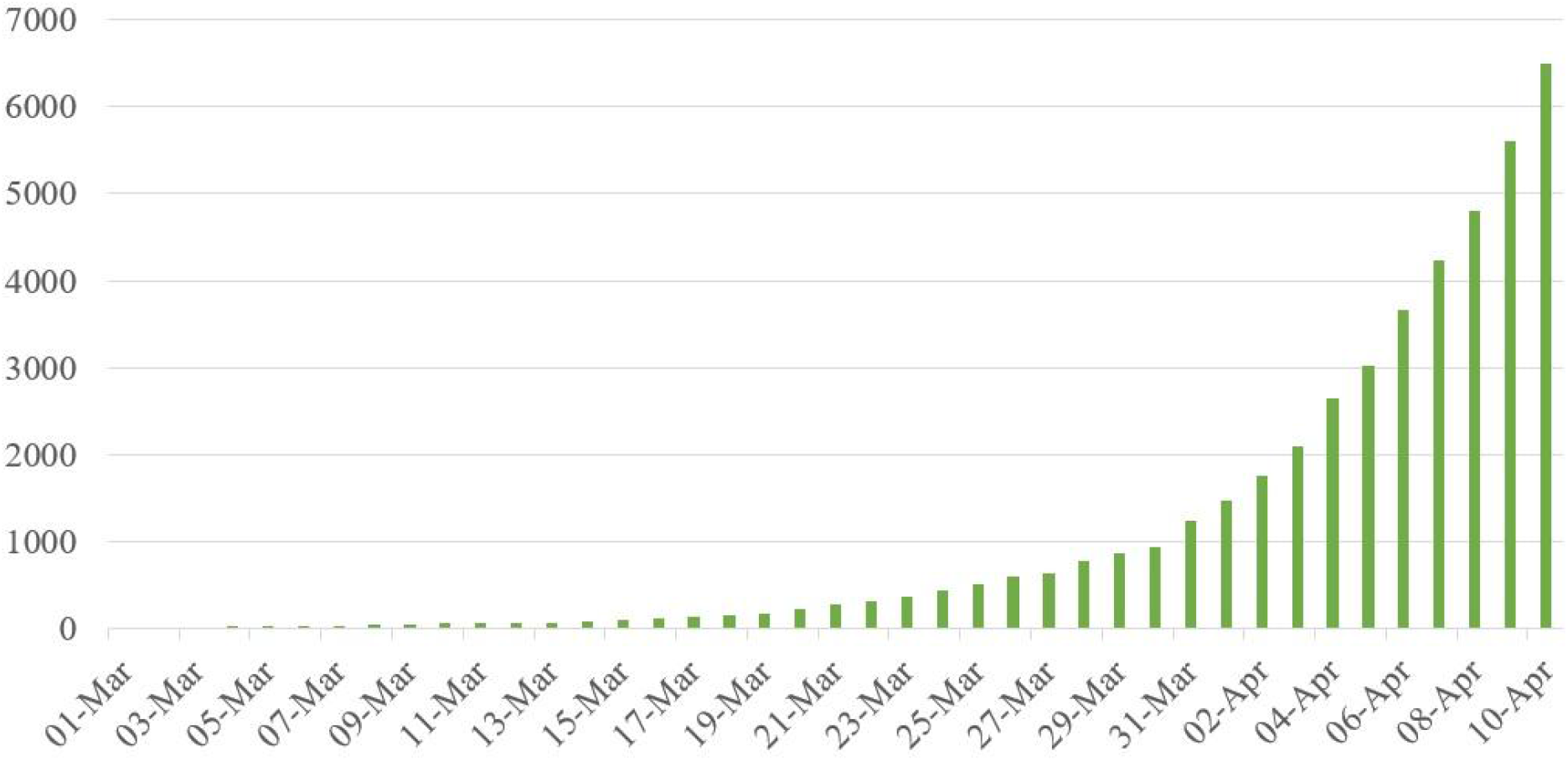
Day-wise active COVID-19 cases in India

The state-wise COVID-19 cases are Tabulated in the Table 1. It can be easily analyzed from the Tab. 1 that Maharashtra, Delhi, Tamil Nadu, Uttar Pradesh, Madhya Pradesh, Telangana and Rajasthan are major affected states with COVID-19 diseases. The COVID-19 spread behavior with different measurements such as before and after applying social distancing and spread due to breakage in maintaining social distancing have been shown in Fig. 3.

**Table 1.** Total number of confirmed state wise cases in India till 17-March-2020

| Name of State/UT | Total confirmed cases | Cured/Discharged | Death |
| --- | --- | --- | --- |
| Andaman and Nicobar Islands | 11 | 10 | 0 |
| Andhra Pradesh | 534 | 20 | 14 |
| Arunachal Pradesh | 1 | 0 | 0 |
| Assam | 35 | 5 | 1 |
| Bihar | 74 | 29 | 1 |
| Chandigarh | 21 | 7 | 0 |
| Chhattisgarh | 33 | 17 | 0 |
| Delhi | 1578 | 42 | 32 |
| Goa | 7 | 5 | 0 |
| Gujarat | 871 | 64 | 36 |
| Haryana | 205 | 43 | 3 |
| Himachal Pradesh | 35 | 16 | 1 |
| Jammu and Kashmir | 300 | 36 | 4 |
| Jharkhand | 28 | 0 | 2 |
| Karnataka | 315 | 82 | 13 |
| Kerala | 388 | 218 | 3 |
| Ladakh | 17 | 10 | 0 |
| Madhya Pradesh | 1120 | 64 | 53 |
| Maharashtra | 2919 | 295 | 187 |
| Manipur | 2 | 1 | 0 |
| Meghalaya | 7 | 0 | 1 |
| Mizoram | 1 | 0 | 0 |
| Nagaland | 0 | 0 | 0 |
| Odisha | 60 | 18 | 1 |
| Puducherry | 7 | 1 | 0 |
| Punjab | 18 | 27 | 13 |
| Rajasthan | 1023 | 147 | 3 |
| Tamil Nadu | 1242 | 118 | 14 |
| Telengana | 698 | 120 | 18 |
| Tripura | 2 | 1 | 0 |
| Uttarakhand | 37 | 9 | 0 |
| Uttar Pradesh | 773 | 68 | 13 |
| West Bengal | 231 | 42 | 7 |

Fig. 3(a) shows polynomial spread behavior with degree 2 before applying social distancing. The effect of social distancing can be seen in Fig. 3(b) which shows a reduction of spread behavior from polynomial to linear. Exponential behavior has been analyzed by breaking the social distancing as can be seen in Fig. 3(c). The overall spread behavior up to 10 April 2020 is shown in Fig. 3(d). Fig. 3(d) shows how spread behavior has been changed from polynomial of degree 2 to polynomial of degree 3. The overall population has been considered as susceptible cases. The experimental studies have been performed on 10000 population. The cases which do not belong to susceptible cases either belong to exposed, infected or recovered. The peoples who have contacted recently to infected people move to exposed cases. The exposed cases move to infected after completion of the incubation period. The infected people's moves recover cases after successful completion of coronavirus cycle, clinical process or death.

**Fig. 3.**
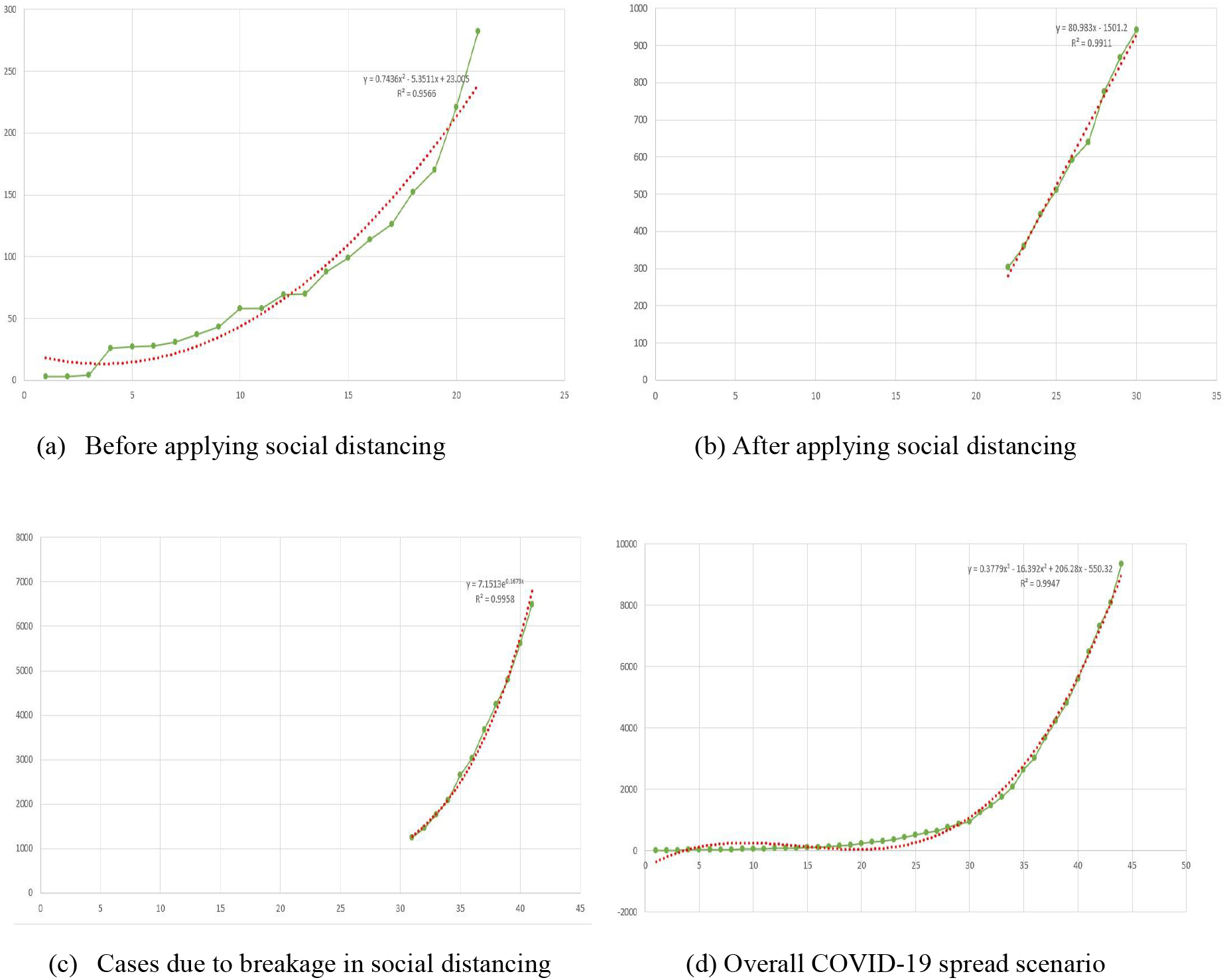
Active COVID-19 cases spread scenarios in India

The value of incubation period α has been fixed with 0.2 and an infection period of 0.5 days has been considered in all modes for validating proposed methodology16, 17. The performance of the proposed mathematical model works in two different modes. The first mode works on the classical SEIR model in which the value of the social distancing rate ρ is fixed to 1. In this mode, the spread of disease depends only on the contact rate β. The result obtained by proposed mathematical model with ρ=1 and β=1.5, β=1.75 and β=2.0 has been shown in the Fig. 4. Green line shows COVID-19 spread with β=1.5, similarly red and blue line shows spread with β =1.75 and β =2.0 respectively. It can be easily analyzed from the green dotted line that spread is about 3% in 21 day, which is approximately the same as COVID-19 disease spread before applying social distancing in India. It can be also analyzed that if this spread has been followed then the maximum 35% population could have been suffered from COVID-19 disease in 30 days.

**Fig. 4.**
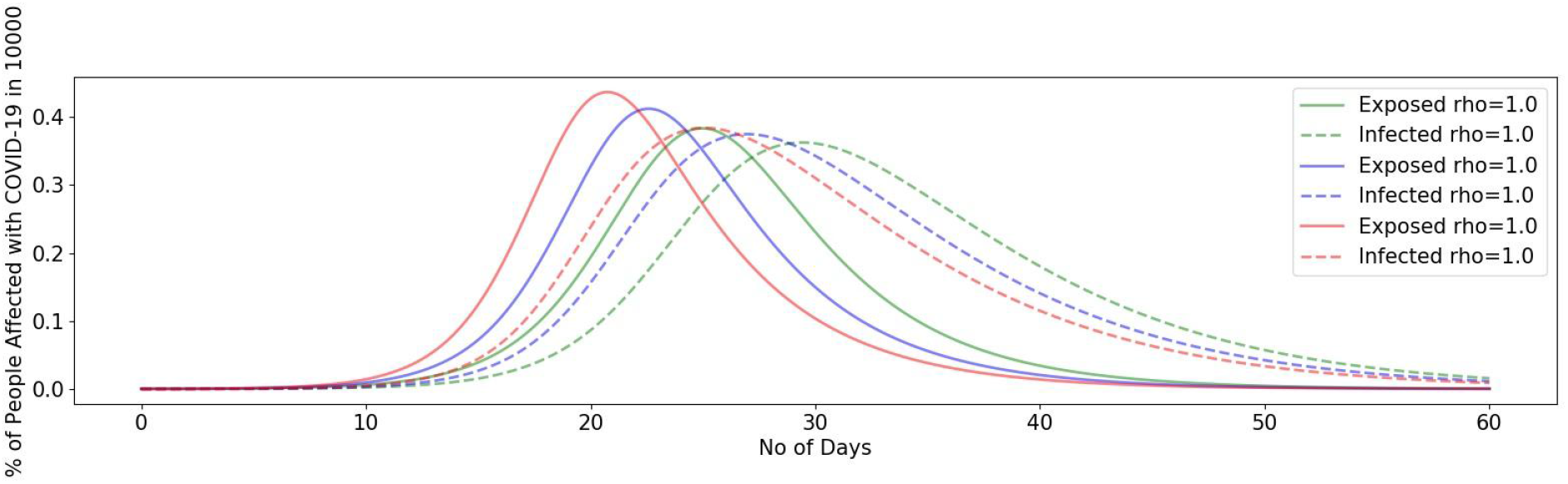
COVID-19 Spread before applying social distancing with ρ = 1

The second mode is with different social distancing rates. The results obtained have been shown in Fig. 5. It has been analyzed with about 300 active cases. It can be easily analyzed from the green dotted line that spread is about 10% in 10 days, which is approximately the same as COVID-19 disease spread after applying social distancing in India. Hence, it can be also analyzed that spread behavior is linear in nature. In addition, it can be said that if this spread has been followed then the maximum 30% population could have been suffered from COVID-19 disease 30 days.

**Fig. 5.**
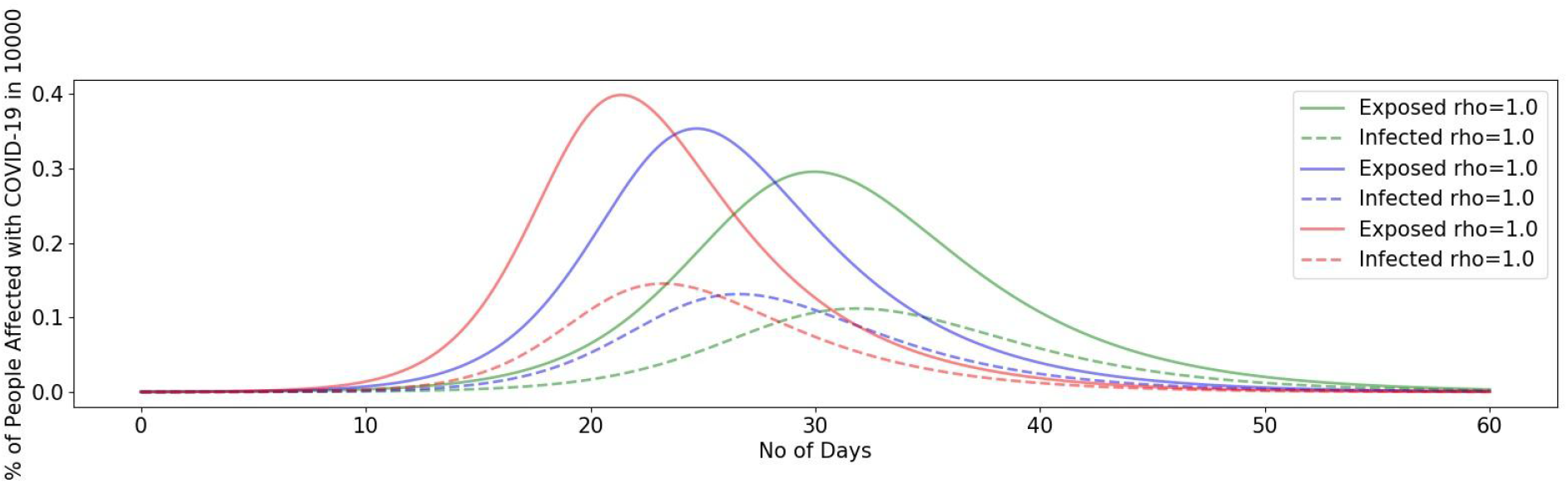
COVID-19 Spread after applying different social distancing rates

The sudden increase has been analyzed after 30 March 2020. This is due to the breakage of social distancing as a gathering of peoples. It can be easily observed from Fig. 6 that spread is about to 60% peoples in 10 days which closely resembles COVID-19 spread after gathering. Hence, it can be said that due to this gathering the spreading behavior has been changed from linear to a polynomial of degree 3. In addition, it is found that 60 days lockdown is required in the current scenario to complete recovery of COVID-19 disease.

**Fig. 6.**
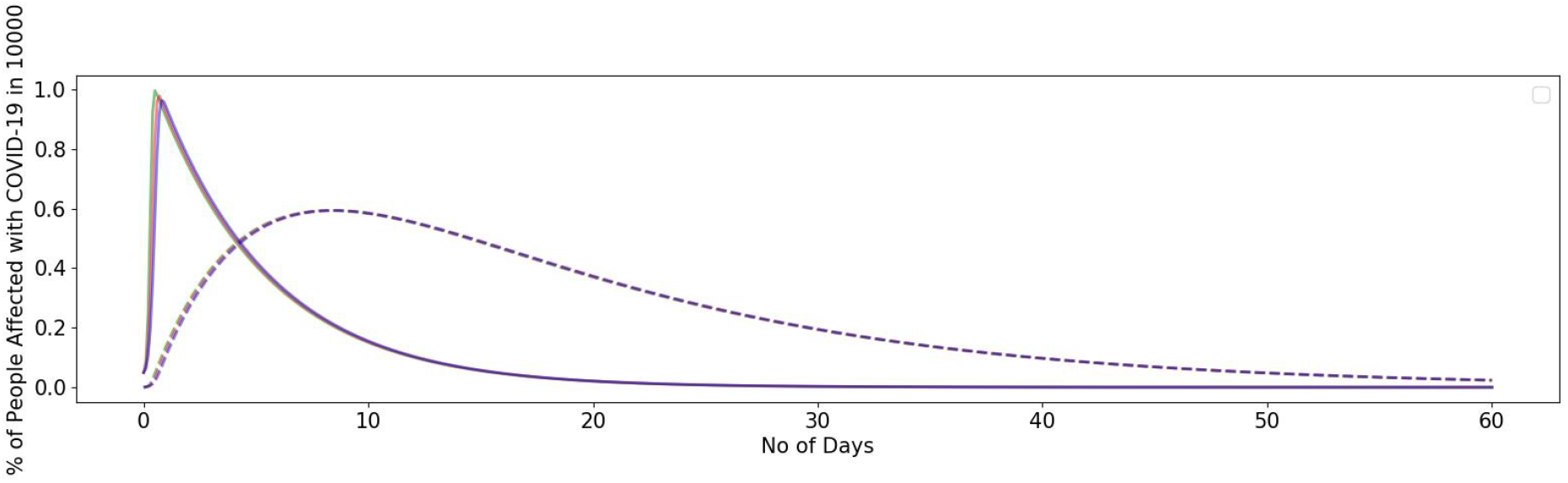
COVID-19 Spread due to breakage in social distancing

## 4. Conclusion

The present work has investigated the problem of COVID-19 spread in India in current scenarios. A mathematical model has been established, which follows the actual data trend of COVID-19 spread in India. It has been proved from analytical (based on mathematical modeling) and simulation results that social distancing plays an important role in spread estimation. The effect of social distancing has been discussed with different social distancing rates. It has been found that social distancing can reduce spread from polynomial to linear. It has been also observed breakage in social distancing can rise spread from linear to exponentially. The maximum cases and recovery periods are also analyzed. It has been found that in the current spread scenario 60 days lockdown is required for complete recovery.

## Data Availability

Code will be available on request

## Acknowledgment

The work has been supported by a grant received from the Ministry of Education, Government of India under the Scheme for the Promotion of Academic and Research Collaboration (SPARC), 2019.

